# MitoFit: Evaluation of a Mitochondrial Fitness Science Communication Intervention for Aging Adults

**DOI:** 10.1101/2024.07.04.24309859

**Authors:** Cathy A. Maxwell, Brandon Grubbs, Mary S. Dietrich, Jeffrey T. Boon, John Dunavan, Kelly J. Knickerbocker, Maulik R. Patel

**Author notes:** **The study was funded by a Vanderbilt University Seeding Success Faculty Award.

## Abstract

A key driver that leads to age-associated decline and chronic disease is mitochondrial dysfunction. Our prior work revealed strong community interest in the concept of *mitochondrial fitness* that led to development of a video based science communication intervention to prompt behavior change in adults aged 50+.

**Aim:** To conduct formative and summative evaluations of *MitoFit,* an instructional, biologically based communication intervention aimed at improving physical activity (PA) in older adults, aged 50+.

**Methods:** *Phase 1 formative evaluation-* Community-dwelling older adults (N=101), rated the acceptability, appropriateness and helpfulness of our MitoFit video series, titled, “How to Slow Down Aging Through Mitochondrial Fitness.” (≥ 4 out of five on a Likert-scale survey). *Phase II summative evaluation*- A subgroup of phase I participants (N=19) participated in a 1-month MitoFit intervention prototype to evaluate intervention and data collection feasibility (≥ 70% completion).

**Results:** *Phase I:* Participants (mean age: 67.8 [SD 8.9]; 75% female) rated the MitoFit videos as acceptable (agree: 97%-100%), appropriate (agree: 100%) and helpful (agree: 95%-100%) to support adaptation and continued work on our novel approach. *Phase II:* Participants (mean age: 71.4 [SD 7.9]; 72% female) demonstrated MitoFit competencies (obtaining pulse, calculating maximum and zone 2 heart rate, demonstration of exercises). At one-month post-instruction, 13 participants (68.4%) had completed a self-initiated daily walking/exercise plan and submitted a daily activity log. Feasibility scores ranged from 89.4% to 94.7%. Fifteen participants (78.9%) stated an intention to continue the MitoFit intervention.

**Conclusion:** MitoFit was enthusiastically embraced, and is a cost-effective, scalable, and efficacious intervention to advance with community-dwelling older adults.

---

Key risk factors for noncommunicable chronic diseases (NCDs) are primarily lifestyle-related with physical inactivity as a leading risk.^1^ Sedentary lifestyles contribute to the top three causes of NCD-related deaths: cardiovascular disease (CVD), type 2 diabetes, and cancer.^2^ In the U.S., only one in four adults meet national physical activity guidelines for aerobic and strength-training activity; with inactivity increasing as individuals age.^3^

A growing body of literature identifies mitochondria as central biological hubs for impaired energy metabolism that leads to development of NCDs.^4, 5^ Mitochondria are essential for maintenance of cellular functions and homeostasis through production of adenosine triphosphate (ATP) for cellular energy, as well as metabolites that support calcium regulation, redox balance, apoptosis and protein synthesis.^6, 7^ Mitochondrial functions modulate multisystemic stress responses, with far-reaching implications for health and disease.^8^ Mitochondrial dysfunctions influence all hallmarks of aging^9^ which is a central focus of the rapidly accelerating field of geroscience. The centrality of mitochondria as a key factor for healthy aging^10^ led to our teams’ interest in advancing understanding among the public-at-large, particularly older adults, as well as health care clinicians.^11, 12^ Our prior work revealed strong community interest in the topic of energy homeostasis; specifically *how the body makes energy*;^12^ and we subsequently developed and tested a health and wellness program, AFRESH, incorporating didactic content on mitochondrial fitness.^13^

To expand this work, we developed a video-based science communication intervention, *MitoFit*, aimed at prompting intrinsic motivation for exercise through enhanced science communication. We wished to explore whether lay-friendly delivery of information about the importance of mitochondria would lead to a change in behavior related to physical activity (PA) in aging adults. Intrinsic motivation entails doing an activity for its inherent meaning vs. external results or rewards.^14^ The reward of intrinsic motivation comes from the behavior itself and the psychological well-being associated with autonomy (need to be in charge) and competence (ability to do).^14^ Systematic reviews on barriers and motivators of PA in older adults report concerns about health/fitness and lack of motivation as key barriers.^15^ Motivators of PA include a person-centered approach and inclusion of behavior capability principles.^16^ Appreciation and respect for scientific information (vs. non-scientific) also influence uptake of health recommendations.^17, 18^

These concepts and principles guided development of our MitoFit video series, titled, “How to Slow Down Aging Through Mitochondrial Fitness,” as well as a prototype MitoFit intervention, a science-based communication intervention targeted to adults aged 50 and older. This paper describes two phases of the MitoFit project: 1) formative evaluation to support acceptability, appropriateness and helpfulness of the MitoFit video series; and 2) summative evaluation to pilot test a MitoFit prototype intervention. Our research questions were: 1) Do community-dwelling adults, aged 50+, find MitoFit videos acceptable, appropriate, and helpful?; and 2) Can the MitoFit prototype intervention and data collection protocol be feasibly implemented with community-dwelling adults over a one-month period? We hypothesized that the MitoFit videos would be acceptable (≥ 4 out of 5 on a Likert scale); the MitoFit prototype intervention would be feasible (≥ 4 out of 5 on a Likert scale); and that our MitoFit approach using science communication and simple instructions would result in *self-initiation* of an exercise routine (≥ 70%) over a one-month period.

## Methods

### Study Design

The study was approved by the Institutional Review Board at Vanderbilt University (IRB #230300). Development and testing of MitoFit was guided by the NIH Stage Model for Behavioral Intervention Development (Figure 1).^19^ The model provides a development roadmap and advances best practices for generating, testing and implementing interventions that can be delivered in real world settings.^20^

**Figure 1.**
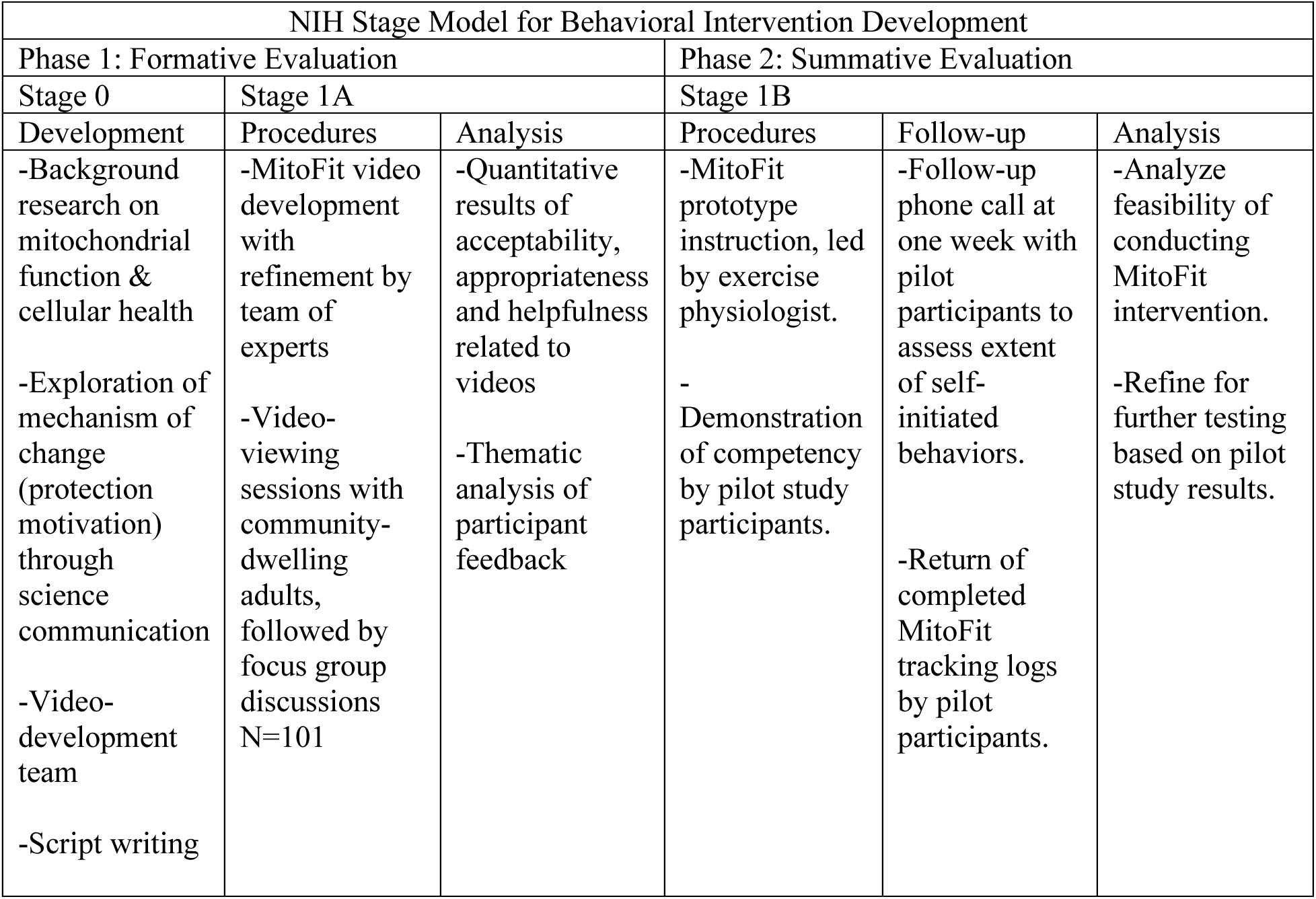
MitoFit Development based on NIH Stage Model: Formative and Summative Phases

### MitoFit Development: Phase 1, Stage 0

Stage 0 of the NIH model involves using the underlying basic science and relevant information for planning prior to intervention development.^20^ During this stage, MitoFit videos were formulated within the domain of social cognitive theory (use of psychology, education, communication) and protection motivation theory of persuasive communication to prompt individuals to engage in behavior to protect themselves.^21, 22^ The MitoFit content and intervention is guided by basic science on mitochondrial function and supported by expert consensus on the use of physical activity for health promotion and disease prevention.^23^ Instruction about physical activity within MitoFit reflects principles of mitochondrial function, cardiorespiratory health (exercise and heart rate zones), and physical exertion levels that translate to improvements in physical fitness (increased physical endurance) and mitochondrial fitness (improved cellular oxygen consumption, VO2max [maximal oxygen consumption]).^24, 25^

### MitoFit Development: Phase 1, Stage 1

Stage 1of the NIH Stage model comprises intervention generation and adaptation (Stage 1A) and pilot testing (Stage 1B).^20^ During this stage, we formed a team comprised of the principal investigator (CAM), a mitochondrial scientist (MP), a kinesiologist/exercise physiologist (BG), experiential learning expert (JD) and graphic designers.

Stage 1A: Over a one-year period (Jan-Dec 2022), our team iteratively developed a series of six short videos (4-6 minutes/each), titled “How to Slow Down Aging Through Mitochondrial Fitness.” Titles and content of individual videos (Table 1) were developed to convey a basic, but simple and practical understanding of the importance of mitochondria for maintaining health and wellness. Our team was guided by literature on science communication to lay audiences.^17, 18^ Scripts were developed by the study team and subsequently incorporated into sequential storyboards (graphic organizers) with suggestions for graphics to accompany specific content. We were mindful of narratives that simplified content and captured attention through messaging that audiences would remember. Audio-recordings to accompany graphic designs were produced in a recording studio with a spokesperson who is shown narrating the videos. Graphic designers combined audio-recordings with graphics, animation and sound to produce individual videos that the team reviewed, revised and approved. Once the videos were developed, phase 1 testing was conducted (see procedures below) to determine acceptability, appropriateness, and helpfulness of the videos. After video development, a MitoFit intervention prototype was developed by the PI and kinesiology researcher with specific expertise in exercise with older adults to accompany the video series (see Table 1).

**Table 1.** Titles of MitoFit Videos and Components of the MitoFit Prototype Intervention.

| Video Series: How to Slow Down Aging through Mitochondrial Fitness | MitoFit Intervention Components |
| --- | --- |
| Video 1: Overview<br>Video 2: What are mitochondria?<br>Video 3: How mitochondria become damaged<br>Video 4: How to create new and better mitochondria<br>Video 5: Keeping our energy requirement in balance<br>Video 6: Making mitochondrial fitness an essential part of daily life | • Calculation of maximum heart rate<br>• Determination of heart rate zone 2<br>• Completion of a one mile walk OR 2000 steps (continuous)<br>• Measurement of heart rate via pulse oximeter<br>• Demonstration of five strength training calisthenics<br>○ Chair stand<br>○ Dumbbell shoulder press<br>○ Wall push-ups<br>○ Calf-raises<br>○ Toe taps |

### Phase 1 Setting and Eligibility

Group sessions were scheduled at five sites (neighborhood [1], senior centers [3], clinical research lab [1]). Participants were recruited via posted flyers at each locale. Eligibility criteria included: English-speaking, age of 50 or older, ability to attend an in-person group session, and ability to walk independently. Eligible participants were offered a $40 gift card upon completion of the group session. Phase 1 participants (N=101) completed written informed consent and were provided copies of the consent.

### Phase 1 Procedures

Sixteen group sessions were scheduled at each of the five sites. Interested individuals either called the PI’s office and left a message or completed an electronic form to indicate interest in the study. The study project coordinator then contacted each person to confirm eligibility and schedule individuals for a group session of up to 12 participants/group. The sessions were led by the project coordinator (14 sessions) or another member of the study team (2 sessions). Each group session began with completion of the informed consent, followed by completion of a demographic form that also included three multiple choice questions to assess baseline knowledge about mitochondria. The group leader then introduced the video series and provided participants with a pen and blank sheet of paper to take notes. To prevent information bias, participants were instructed to hold their questions until all six videos were viewed and survey questions completed. Videos were viewed by the groups, two videos at time, pausing in between for a focus group discussion. The focus groups were led by a group leader and included four open-ended questions with probes:

#### Qualitative questions

- What is your initial reaction to the videos using one or two words?
- What do you think is the most important take-way from the videos?

`o` What new information did you learn?
`o` How did the videos help in understanding mitochondrial fitness?
- What changes, if any, would you suggest to make the videos more effective?
- What was the most memorable part of the videos for you?

`o` How did the videos improve your understanding of mitochondrial fitness?
`o` How could you use the information from the videos in your day-to-day life?

The focus group discussions (3 per group session) were audio-recorded; and the mp3 files were subsequently uploaded to a transcription service (Rev.com) and transcribed verbatim. Given the quantity and relevance of qualitative data from the focus groups, the authors developed a second publication that is under peer review in another journal. Upon completion of the focus groups, participants completed a 15-item survey on acceptability, appropriateness, and helpfulness (see data collection below). At the end of the survey, participants could provide their phone number to receive a call in one week for us to inquire about perceptions after participants had time to reflect more on the videos. After all data collection was completed, the PI offered to answer any questions that the participants had about the videos.

### Phase 1 Data Collection

Phase 1 participants (video viewers) completed a demographic form that included age, race/ethnicity, gender, marital status, education level, employment status and three questions about mitochondria. Questions for acceptability, appropriateness, and helpfulness were collected via surveys adapted from the Acceptability of Intervention Measure (AIM) and Intervention Appropriateness Measure (IAM)^19^ and responses reflected agreement (completely disagree to completely agree) on a 5-point Likert scale. Data were collected via paper surveys and double-entered/coded in REDCap, an encrypted, secure electronic data collection system.^20, 21^

### MitoFit Development: Phase 2, Stage 1B

During this summative phase of the NIH Stage Model, we assessed the feasibility of administering a prototype MitoFit intervention that would accompany the videos and the data collection processes planned for use with community-dwelling adults, aged 50+. The MitoFit intervention protocol entailed: a) viewing the MitoFit videos (simplified science communication on mitochondria), b) didactic instruction about heart rate zones to create cellular demand for energy, c) demonstration of five competencies (see Table 1), and d) guidance on how to engage in a self-initiated walking and exercise plan. We proposed that our approach (science communication + simple instructions) would lead to self-initiation of a walking and strength training routine over a one-month period. Age 50 was chosen for eligibility because our prior study revealed the 45-65 age group having the strongest likelihood to change behavior.^12^ For phase 2 pilot testing of the prototype intervention, a subgroup of phase 1 participants (N=19) volunteered to receive the MitoFit intervention. Testing of the video series and MitoFit intervention was conducted over 8 months (May-December 2023).

### Phase 2 Prototype Development

Development of the MitoFit intervention prototype was based on evidence that individuals are more likely to self-initiate exercise routines that are simple, doable, and efficacious, with the ability to do the intervention within one’s own home or vicinity.^22^ A central message of the intervention content is that *mitochondrial health and energy homeostasis is essential to healthy aging, and improved/maintained by creating the demand for energy through regular sustained physical activity.* The intervention is based on physical activity guidelines for Americans^23^ and includes information about activities that comprise moderate and vigorous physical activity based on heart rate zones and physical exertion.^24^ We specifically promoted moderate activity performed at heart rate zones 1 and 2 (50% to 70% of the maximum heart rate).

### Phase 2 Eligibility

Participation in phase 2 was voluntary and offered to phase 1 participants. Eligibility included: viewing of the MitoFit videos, absence of heart failure and the ability to walk independently for 20 minutes. Use of a cane or other walking aid was permitted. Participants were offered an additional $40 gift card upon completion of the group 2 session.

### Phase 2 Procedures

After completion of a second informed consent, the phase 2 group sessions began with completion of surveys for baseline physical activity and quality of life (see data collection below), followed by didactic instruction. Sessions were led by a certified exercise physiologist/kinesiology researcher (BG). Participants were instructed on heart rate zones and provided rationale and illustrations about how heart rate reflects utilization of nutrients (substrates: fatty acids or glucose) by mitochondria for fuel and energy. We promoted sustained/continuous physical activity in heart rate zones 1 and 2 for at least 20-30 minutes, emphasizing that these zones are typically perceived as doable at an exertion level in which participants can engage in conversation while walking. We also taught participants how to complete five basic calisthenic exercises, including chair stands, dumbbell shoulder press, wall push-ups, calf-raises, and toe taps. Participants were provided with a pulse oximeter (finger) and step counter that clipped to their clothing. Upon completion of didactic instruction, we explained how to estimate the age-based maximum heart rate and heart rate zone ranges for each participant using the following formula: 207 – (0.7 X age) = maximum heart rate.^25^ We then had participants demonstrate competencies: 1) calculation of their maximum heart rate, 2) determination of zones 1 & 2 heart rates, 3) obtaining their pulse via a pulse oximeter, and 4) and completion of five calisthenic exercises. To conclude, participants were given a daily exercise tracking log and asked to record their initial walking pulse in heart rate zones 1 or 2, minutes walked in one mile, days of walking over one month, miles walked each day, and strength training over one month. We told the participants that we would call them in 1 week to check in and inquire if they had initiated their personal plan, and again at one-month to ask them to send us a copy of their completed exercise tracking log.

### Phase 2 Data Collection

Phase 2 participants completed the Rapid Assessment of Physical Activity (RAPA; 9-items [7 items-aerobic, 2 items- strength/flexibility])^26^ and the Patient-Reported Outcomes Measurement Information System (PROMIS-19; 19 items)^27^ that assesses quality of life in 7 domains (physical function, anxiety, depression, fatigue, sleep, socialization, pain). Upon completion of the group 2 session, participants completed the Feasibility of Intervention Measure (FIM).^28^ At one week, we called the participants to ask if they had self-initiated a walking and strength training plan and if they had used their pulse oximeter to monitor their heart rate when they walked. At one-month after the group 2 session, we called participants and readministered the RAPA, as well as the three questions about mitochondria that we had asked during the original group 1 sessions. We were interested if the participants retained knowledge about mitochondrial fitness. Finally, at one-month post group 2, we asked participants to send us (via email or text) a copy of their completed exercise tracking log. Collected data from paper sources were transferred into REDCap.

### Phase 1 and 2 Data Analysis

Data analysis was conducted using IBM SPSS Statistics (Version 29.0). We used frequency distributions to summarize most of the data collected in both phases of the study. Mean (SD) were used for the continuous age distributions and median (IQR) were used for the PROMIS-19, RAPA, and pain ratings. Changes in the RAPA values (Phase 2) were analyzed using Wilcoxon Signed-Ranks tests. Any interpretation of statistical significance used a critical alpha value of .05 (i.e., p < .05).

## Results

### Phase 1 Formative Evaluation

One hundred and one participants attended one of the 16 group sessions. A majority (n=74, 73.3%) were between the ages of 60 and 80 years and 27.3% (n=27) identified as African American. Approximately half (53%, n=53) had a college degree or higher, 75% (n=75) identified as female, almost half (47.5%, n=48) were married or partnered; and 61.4% (n=62) were retired. Prior to viewing the videos, participants’ correct responses to the mitochondrial knowledge questions ranged from 51.5% (What are mitochondria?) to 80.4% (What is the primary function of mitochondria?). (Table 2)

**Table 2.**
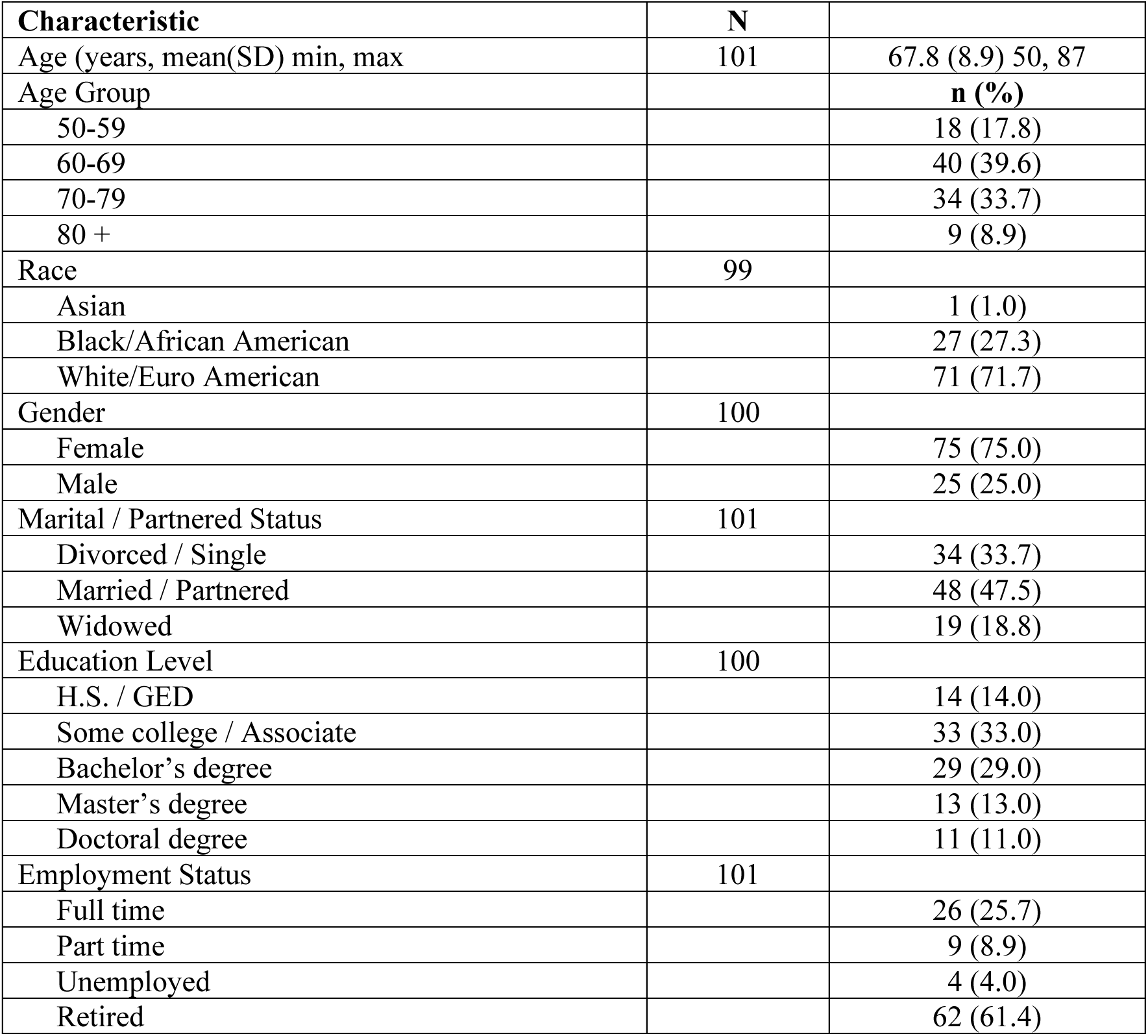
Characteristics of Study Participants and Knowledge of Mitochondria (N=101)

### Acceptability, Appropriateness, and Helpfulness of Videos

Agreement scores (4 or 5) for acceptability, appropriateness and helpfulness ranged from 94.1% to 100% with highest scores for appropriateness (Table 3). To explore potential intrinsic motivation related to the videos, we contacted 49 (48.5%) group 1 participants at one week who provided their phone numbers and asked, “Since you viewed the MitoFit videos, have you begun any physical activity that you were not already doing?” Thirty-five (71.4%) stated that they had self-initiated walking or other exercise (e.g., group class).

**Table 3.** MitoFit Videos Acceptability, Appropriateness and Helpfulness (N=101)

|  | N |  |
| --- | --- | --- |
| **MitoFit Video Evaluation** | 101<sup>a</sup> | n (%) [Agree] |
| **Acceptability** |  |  |
| Content engaging |  | 100 (99.0) |
| Length appropriate |  | 97 (96.0) |
| Information easy to understand |  | 97 (96.0) |
| Videos meet my approval |  | 97 (96.0) |
| Videos are appealing to me |  | 98 (97.0) |
| Like the videos |  | 98 (97.0) |
| Welcome the videos | 99 | 98 (99.0) |
| **Appropriateness** |  |  |
| Seem fitting for people aged > 50 |  | 101 (100.0) |
| Seem suitable for people aged > 50 |  | 101 (100.0) |
| Seem applicable for people aged > 50 | 100 | 100 (100.0) |
| Seem like good match for people aged > 50 |  | 101 (100.0) |
| **Helpfulness** |  |  |
| Increased my understanding of mitochondrial fitness |  | 99 (98.0) |
| Would be positive addition for older adults |  | 100 (99.0) |
| Motivated me to want to exercise more | 100 | 95 (95.0) |
| Feel more in control of my health and aging |  | 95 (94.1) |
| **Self-initiated behavior change (one week post-phase 1)** | 49 |  |
| No |  | 14 (28.6) |
| Yes |  | 35 (71.4) |
| **Knowledge of Mitochondria** |  | **n (%) Correct** |
| What are mitochondria? | 97 | 50 (51.5) |
| What is the primary function of mitochondria? | 97 | 78 (80.4) |
| The best way to maintain mitochondrial fitness is to: | 98 | 59 (60.2) |

<sup>a</sup> Unless noted otherwise below, the total number respondents to the respective question was 101.
<sup>b</sup> A response of 4 or 5 on a Likert-response scale ranging from 1 to 5.

### Phase 2 Summative Evaluation

Nineteen participants who confirmed that they met eligibility criteria volunteered to attend a group 2 session to receive the MitoFit prototype intervention. As shown in Table 4, their demographic characteristics were very similar to the entire group participating in Phase 1.

**Table 4.** Characteristics of Group 2 MitoFit Intervention Participants (N=19)

| Characteristic | N |  |
| --- | --- | --- |
| Age (years, mean (SD) min, max | 19 | 71.4 (7.9) 54, 86 |
| Age Group |  | **n (%)** |
| 50-59 |  | 1 (5.3) |
| 60-69 |  | 5 (36.8) |
| 70-79 |  | 8 (42.1) |
| 80 + |  | 3 (15.8) |
| Race | 17 |  |
| Asian |  | 1 (5.9) |
| Black/African American |  | 1 (5.9) |
| White/European American |  | 15 (88.2) |
| Gender | 18 |  |
| Female |  | 13 (72.2) |
| Male |  | 5 (27.8) |
| Marital / Partnered Status | 19 |  |
| Divorced / Single |  | 2 (10.5) |
| Married / Partnered |  | 11 (57.9) |
| Widowed |  | 6 (31.6) |
| Education Level | 19 |  |
| H.S. / GED |  | 1 (5.3) |
| Some college / Associate |  | 8 (42.1) |
| Bachelor's degree |  | 7 (36.8) |
| Master's degree |  | 3 (15.8) |
| Doctoral degree |  | 0 (0.0) |
| Employment Status | 19 |  |
| Full time |  | 2 (10.5) |
| Part time |  | 1 (5.3) |
| Retired |  | 16 (84.2) |
| PROMIS-19 T-scores | 19 | Median(IQR) min, max |
| Physical function (Higher is better) |  | 57.0 (42, 57) 34, 57 |
| Participate social activities (Higher is better) |  | 55.5 (50, 65) 37, 65 |
| Anxiety (Lower is better) |  | 55.3 (40, 58) 40, 61 |
| Depression (Lower is better) |  | 41.0 (41, 56) 41, 64 |
| Fatigue (Lower is better) |  | 48.6 (33, 54) 33, 62 |
| Sleep disturbance (Lower is better) |  | 49.6 (46, 54) 32, 59 |
| Pain interference (Lower is better) |  | 52.4 (41, 56) 41, 67 |
| Average rating of pain (0-10) | 19 | 2.0 (0, 5) 0, 8 |

Among 7 domains related to quality of life, median T-scores were better than the national average for U.S. adults in physical function, depression, fatigue, sleep disturbance and social activities, and worse than average for anxiety and pain interference. The median pain rating was 2.0 on the 0-10 scale (IQR: 0, 5).

### MitoFit Feasibility

The feasibility ratings in the “agree/completely” categories for the intervention prototype ranged from 89.4% (seems easy to follow) to 94.7% (implementable, possible, doable) (Table 5). Among the four competencies of the intervention protocol, all but one participant was able to demonstrate competence. The same participant was unable to calculate a maximum heart rate and heart rate zones. Upon completion of intervention training, 18 of 19 participants agreed (score: 4 or 5) that the intervention was implementable, possible, and doable; and 17 agreed that the intervention was easy to follow.

**Table 5.** MitoFit Prototype Intervention Feasibility (N=19)

|  |  |  |
| --- | --- | --- |
| Table 1. Number of Participants per Intervention Feasibility (n = 19) |  | n (%) |
| Feasibility |  |  |
| The intervention seems implementable. |  |  |
| Disagree/Completely disagree |  | 1 (5.3) |
| Neither Agree/Disagree |  | 0 (0.0) |
| Agree/Completely agree |  | 18 (94.7) |
| The intervention seems possible. |  |  |
| Disagree/Completely disagree |  | 1 (5.3) |
| Neither Agree/Disagree |  | 0 (0.0) |
| Agree/Completely agree |  | 18 (94.7) |
| The intervention seems doable. |  |  |
| Disagree/Completely disagree |  | 1 (5.3) |
| Neither Agree/Disagree |  | 0 (0.0) |
| Agree/Completely agree |  | 18 (94.7) |
| The intervention seems easy to follow. |  |  |
| Disagree/Completely disagree |  | 1 (5.3) |
| Neither Agree/Disagree |  | 1 (5.3) |
| Agree/Completely agree |  | 17 (89.4) |
|  | Baseline | 4-Weeks |
|  | n (%) Correct | n (%) Correct |
| Knowledge of Mitochondria |  |  |
| What are mitochondria? | 14 (73.4) | 12 (63.2) |
| What is the primary function of mitochondria? | 18 (94.7) | 19 (100.0) |
| The best way to maintain mitochondrial fitness is to: | 10 (52.6) | 19 (100.0) |
|  | n (%) |  |
| Completed/submitted MitoFit tracking form | 13 (68.4) |  |
| Intend to continue MitoFit walking/exercise plan | 15 (78.9) |  |

At one-month post-intervention delivery, 15 participants (78.9%) stated that they had implemented a walking and exercise plan, describing what they had done over the month. Thirteen (68.4%) participants completed and submitted a self-initiated daily activity log (see supplemental material S3). Within the group of Phase 2 participants, knowledge of mitochondria improved to 100% from their Phase 1 responses for two of the three questions (primary function and how to maintain) (Table 5).

Self-reports from the 19 participants in Phase 2, indicated a statistically significant increase in aerobic/endurance scores on the RAPA surveys from baseline (Phase 1) to four weeks post-intervention (Phase 2). None of the participants achieved “active” status (30 minutes of moderate activity, 5 days per week), however the percentage of participants in the “under active regular activities” in Phase 1 (35.3%) to 58.8% in Phase 2. While not statistically significant (p = .64), some improvement was noted in strength training, but not in flexibility.

Among the 19 intervention participants, one dropped out at one week before starting a walking plan and one could not be reached for the one-month follow-up despite repeated attempts. We were able to contact 17 participants for follow-up at one-month post intervention. Among all intervention participants, 15 of 19 (78.9%) expressed a stated intention to continue a walking plan. Thirteen (68.4%) participants submitted copies of their exercise tracking logs (jpeg or pdf files) either by email or by phone text. Four participants were contacted but did not submit their tracking logs. Two of the four stated that they tried to submit but were unsure of how. From the one-month follow-up phone calls, An example of a blank tracking log is provided (supplemental material S2).

### Submitted tracking logs

Among the 13 participants who submitted their tracking logs, ages ranged from 64 to 81 (Supplemental material S3). Eleven of 13 utilized their pulse oximeter to obtain a walking pulse in zones 1 or 2 (range: 62-120). All 13 submitters documented the number of minutes walked and the days (dates) walked. The mean number of days walked per month was 21.2 days, ranging from 10 to 31 days. Minutes walked per day ranged from five to 74 minutes. All 13 submitting participants documented the days (dates) that they did the strength training calisthenic exercises. Ten of 13 engaged in strength training exercises, ranging from two to 31 days/month over the one-month period. Three participants did not engage in strength training exercises. At the bottom of the tracking form, participants could fill in weekly totals and eight participants completed the section. One participant documented COVID-19 as the cause for nine missed days. One participant documented the location of their daily walks (i.e., senior center, treadmill, neighborhood).

## Discussion

Formative and summative evaluation of MitoFit demonstrated acceptability, appropriateness and helpfulness of our video series on mitochondrial fitness targeted to adults aged 50 and older. Feasibility of the MitoFit intervention prototype was demonstrated through FIM survey results, demonstration of competencies, self-initiation of personal walking plans (78.9%), and submission of completed exercise tracking logs (68.4%). Additionally, qualitative data from the group 1 focus groups (reported in a separate publication) demonstrated evidence of cognitive restructuring (mental adjustments to prior unhelpful beliefs), influenced by science communication in the videos, didactic content, and illustrations. Comments from group 2 participants during one-month follow-up phone calls reflected change in how individuals regard the importance of exercise for healthy aging. For example, participant #7 stated, “Thank you for the opportunity to be a part of something that has forever changed how I view life and health.” Another participant (#81) stated, “Thank you for doing this study! It has really motivated me to keep it up and it’s not too hard to do.”

From a public perspective, development of the MitoFit videos and intervention prototype was prompted by our team’s awareness that obesity and rising rates of chronic disease in the U.S. constitute a public health crisis;^35^ and that strategies to address this crisis should be efficacious, cost-effective, and scalable to reach large numbers of people. Interventions to increase physical activity in adults abound in scholarly literature, highlighting high costs and difficulty in sustaining behavior change that will truly alter health outcomes. Azjen’s Theory of Planned Behavior^36^ posits that beliefs provide the basis for intention to engage in specific behaviors.

Science communication can influence beliefs and intention.^17, 18^ The MitoFit videos and intervention were developed to prompt protection motivation by increasing knowledge about the importance of mitochondria as essential biological hubs that play a major role in metabolic health and energy homeostasis. We also desired that MitoFit content would impress participants with a sense that daily walking and exercise could be simple, doable and sustainable. We achieved our aims for this pilot work and will utilize study findings to support expansion of MitoFit to larger audiences using cost effective implementation methods (e.g., online learning platform, digital twin technology, public health outreach).

Understanding the role of mitochondria in health and disease will likely grow exponentially over the next decade. Our team supports advancing understanding of mitochondrial fitness among health care providers, equipping them to engage in dialogue with patients to promote proactive decision-making and preventive medicine. Directing patients to lay-friendly sources of publicly-available information on mitochondrial health could be pivotal in turning the tide of obesity and chronic disease in the U.S. and other countries. Geroscience is a new field of medicine that combines aging biology, chronic disease, and health to understand how geroscience-guided interventions, including behavioral interventions like MitoFit, affect progression of chronic disease.^37^ Mitochondrial health intersects with every hallmark of aging and is the foundation of metabolic programming at the cellular level. Aging is characterized by a shift from catabolism to anabolism and from cellular energy production to an emphasis on repair processes as tissue- and organ-level processes begin to breakdown. Explaining these complex mechanisms through science communication to both health care and lay audiences is imperative to address our health care challenges.

### Strengths and Limitations

Strengths of our study include our novel science communication approach for prompting behavior change, as well as a diverse sample of participants for assessing acceptability of the MitoFit videos. Our study achieved promising results through a low-cost communication approach. Our cost-effective approach to intervention delivery and team involvement over the one-month post-intervention period could also be viewed as a limitation since we had minimal engagement with participants during the one-month post group 2 period. Our approach was intentional to enable assessment of whether participants would *self-initiate* behavior change based primarily on communication strategies to prompt planned behavior, a crucial component of motivation and behavior sustainability. We acknowledge participants’ expectation of a follow-up one-week and one-month phone call may have also contributed to motivation. Quantifying motivation for change would strengthen our approach in future studies. Another potential limitation of the intervention prototype testing was a disproportionate percentage of adults aged 70 or older (57.9%) which could have contributed to greater physical limitations, as well as lower digital literacy as reflected in the inability to submit tracking logs via email or text. As adults age, they are less likely to engage in enough PA to cause mitochondrial adaptations.

Future work will include a broader sample of age groups. Other limitations include the short (one-month) follow-up period and omission of flexibility training instruction (a component of the RAPA). Future studies with MitoFit should include a one- or two-year follow-up to determine true sustainability and attrition and inclusion of flexibility exercises.

## Conclusion

Mitochondrial health is a critical mediator of cellular function and metabolic programming that translates to overall health and wellness. These biological processes are complex, however, communication to lay audiences and health care providers will be paramount in the future to promote proactive and preventive behavior change. Communication strategies and interventions like MitoFit that promote physical activity (walking) are simple and doable, with the potential to induce beneficial mitochondrial adaptations that could translate to improvements in health and prevention of chronic disease in aging adults.^37^

## Data Availability

All data produced in the present study are available upon reasonable request to the authors.

## Acknowledgements

Our MitoFit team is appreciative of the Vanderbilt University’s Faculty Seeding Success Award that funded the MitoFit project. The team is also grateful to Sandra Gilbert for serving as our project manager.

