## Supplemental data tables for "MitoFit: Evaluation of a Mitochondrial Fitness Science Communication Intervention for Aging Adults"

Supplemental material S1. Baseline Physical Activity (RAPA) and Demonstration of Competencies (N=19)

| **Rapid Assessment of Physical Activity (RAPA, N=17)** | **Baseline** | **4-Weeks** | **p-value** |
|---|---|---|---|
| Aerobic | | | .028 |
| Sedentary | 1 (5.9) | 0 (0.0) | |
| Under-active | 3 (17.6) | 1 (5.9) | |
| Under-active, regular light activities | 7 (41.2) | 6 (35.3) | |
| Under-active, regular moderate activities | 6 (35.3) | 10 (58.8) | |
| Active | | | |
| Strength and Flexibility | | | .64 |
| None | 3 (17.6) | 4 (23.5) | |
| Strength | 3 (17.6) | 6 (35.3) | |
| Flexibility | 6 (35.3) | 0 (0.0) | |
| Both strength & flexibility | 5 (29.4) | 7 (41.2) | |
| Demonstration of MitoFit competencies | | n (%) | |
| Obtains pulse via pulse oximeter | | | |
| Unable to perform | | 0 (0.0) | |
| Yes, with more than 2 attempts | | 0 (0.0) | |
| Yes, with 2 attempts | | 1 (5.3) | |
| Yes, after 1st attempt | | 18 (94.7) | |
| Calculates maximum heart rate | | | |
| Unable to perform | | 1 (5.3) | |
| Yes, with more than 2 attempts | | 0 (0.0) | |
| Yes, with 2 attempts | | 2 (10.5) | |
| Yes, after 1st attempt | | 16 (84.2) | |
| Calculates zone 2 heart rate | | | |
| Unable to perform | | 1 (5.3) | |
| Yes, with more than 2 attempts | | 0 (0.0) | |
| Yes, with 2 attempts | | 2 (10.5) | |
| Yes, after 1st attempt | | 16 (84.2) | |
| Demonstrates five strength training exercises | | | |
| Unable to perform | | 0 (0.0) | |
| Yes, with more than 2 attempts | | 0 (0.0) | |
| Yes, with 2 attempts | | 2 (10.5) | |
| Yes, after 1st attempt | | 17 (89.5) | |

Supplemental material S2. Example of blank MitoFit Tracking form

**TRACKING MY WALKING AND STRENGTH TRAINING**

| DATE | ACTIVITY | MINUTES | ACTIVITY | MINUTES | VO2MAX |
|---|---|---|---|---|---|
| 1 | | | | | |
| 2 | | | | | |
| 3 | | | | | |
| 4 | | | | | |
| 5 | | | | | |
| 6 | | | | | |
| 7 | | | | | |
| 8 | | | | | |
| 9 | | | | | |
| 10 | | | | | |
| 11 | | | | | |
| 12 | | | | | |
| 13 | | | | | |
| 14 | | | | | |
| 15 | | | | | |
| 16 | | | | | |
| 17 | | | | | |
| 18 | | | | | |
| 19 | | | | | |
| 20 | | | | | |
| 21 | | | | | |
| 22 | | | | | |
| 23 | | | | | |
| 24 | | | | | |
| 25 | | | | | |
| 26 | | | | | |
| 27 | | | | | |
| 28 | | | | | |
| 29 | | | | | |
| 30 | | | | | |
| 31 | | | | | |

Walking/Jogging - Minutes/Week

Week 1: ___________ Week 2: __________ Week 3: __________ Week 4: __________

Strength Training - Minutes/Week

Week 1: ___________ Week 2: __________ Week 3: __________ Week 4: __________

Supplemental Material S3. Summary of submitted tracking log data (by chronological age) at one-month post group 2 session (N=13)

| Participants | Age Group | Walking pulse | Minutes/mile | # Days Walked | # Miles/ Month | # Days Strength Training |
|---|---|---|---|---|---|---|
| ID | 60-64 | 98 | 21.0 | 10 | 10 | 0 |
| ID | 60-64 | 92 | 23.0 | 20 | 20 | 10 |
| ID | 60-64 | 118 | 17.38 | 15 | 18 | 7 |
| ID | 65-69 | 98 | 25.0 | 10 | 10 | 8 |
| ID | 65-69 | 105 | 18.0 | 29 | 29 | 6 |
| ID | 65-69 | 98 | 23.3 | 11 | 16.5 | 2 |
| ID | 70-74 | 104 | 22.0 | 29 | 29 | 12 |
| ID | 70-74 | 120 | 27.08 | 25 | ND | 6 |
| ID | 75-79 | 112 | 20.0 | 31 | 31 | 31 |
| ID | 75-79 | 128 | 20.16 | 31 | ND | 11 |
| ID | 75-79 | - | - | 21 | ND | 0 |
| ID | 80-84 | 62 | 22.0 | 31 | ND | 10 |
| ID | 80-84 | - | - | 12 | ND | 0 |
